# Patients with benign breast disease and breast cancer need more COVID-19 vaccines

**DOI:** 10.1101/2021.12.09.21267531

**Authors:** Huai-liang Wu, Zong-lin He, Yue Gong, Miao Mo, Guang-yu Liu

**Author notes:** These authors contributed equally (Huai-liang Wu, Zong-lin He). Corresponding author **Correspondence** Prof Guang-yu Liu, Department of Breast Surgery, Fudan University Shanghai Cancer Centre, 270 Dongan Road, Xuhui District, Shanghai, 200032, P.R. China.

## Abstract

Albeit the efficacy of COVID-19 vaccines in immunocompromised patients is undermined, it is still found beneficial. Patients with cancer have a much lower COVID-19 vaccination rate globally, and the vaccination coverage in breast cancer patients in China remains elusive. A total of 23029 patients with benign breast diseases and breast cancers were included in the study, and the vaccination rates of patients with benign breast tumors and other benign breast diseases, nonmetastatic and metastatic breast cancer were 44.0%, 54.7%, 19.2% and 9.6%, respectively. Breast cancer in situ patients had a similar vaccination rate with patients with benign breast tumors (45.9% vs 44.0%) while those with invasive breast cancer had much lower vaccination rates. The overall vaccination rate remains meager in breast cancer patients, and gap was found in patients with lower clinical stage. Hence vaccination should be further promoted among patients with benign breast diseases and breast cancer.

## Introduction

During the COVID-19 pandemic, the management of cancer patients has been unavoidably affected.[1] On the one hand, cancer patients are more susceptible to COVID-19 infection due to their immunosuppressive states;[2] on the other hand, the ubiquitous COVID-19-related public health measures render it difficult for their medical seeking, and malpractice of personal protective equipment may inversely increase the risks for contracting infection.[3]

Clinical trials had indicated the effectiveness of COVID-19 vaccines in curbing the outbreak.[4] Although various variants of the SARS-CoV-2 like Omicron and Delta emerged unprecedentedly, the COVID-19 vaccines remain the leading solutions to building the herd immunity against the pandemics. Nevertheless, regardless of the herd immunity, patients with active immunity are still susceptible to high morbidity and mortality of COVID-19, especially for cancer patients, whose immunity is left impaired after sequence of systemic treatments. The immunosuppression secondary to cancer and cancer treatment renders the cancer patients vulnerable to infection, even upon vaccination. Only few studies reported the vaccination uptake in cancer populations, let alone in breast cancer (BC) patients, one of the largest proportions of cancer patients.

Against the prevailing “Co-existing strategies of epidemic control”, China had made considerable efforts in mitigating COVID-19 epidemics by strict and comprehensive regulatory resorts and now entered the normalized regulatory stage.[5] Albeit the increasingly high vaccination coverage rate in China and the strict normalized regulatory measures, the COVID-19 vaccination in BC patients remains elusive and is worth investigating.

## Methods

From August 1 to October 31, 2021, any patients with complete clinical data visiting the Breast Surgery Clinic in the Fudan University Shanghai Cancer Center, Shanghai, China, were included in this study. To assess the relationship between COVID-19 vaccination and breast diseases patients, we matched their epidemiological surveys of COVID-19 vaccines with medical records in the clinic. Diagnosis of benign breast tumor or malignant BC required biopsies. Statistical analysis was performed with IBM SPSS 20.0 software. A two-sided p-value of 0.05 was defined as significant.

## Results

A total of 23029 patients were included in the study, of which 2618 confirmed benign breast tumors, 9308 with other benign breast diseases, 10666 with nonmetastatic BC, and 437 metastatic BC. The vaccination rates of patients with benign breast tumors and other benign breast diseases, nonmetastatic and metastatic BC were 44.0%, 54.7%, 19.2% and 9.6%, respectively (Figure 1). For 11103 diagnosed BC patients, only 2137 (19.2%) BC patients had received COVID-19 vaccines, while 9124 had not upon data collection (Table 1). Among breast cancer patients, breast carcinoma in situ had the highest vaccination rate (45.9%), which was comparable with the vaccination rate of patients with benign breast tumors (44.0%). Besides, patients with lower tumor burden (such as N0 and N1 stage) had significantly higher vaccination rates than patients in cN2 and cN3 stages (21.0% and 17.1% vs 12.3% and 11.7%, p<0.001). Overall, there significant difference of vaccination rates among different clinical stages breast cancer patients (Stage 0, stage I, stage II, stage III and stage IV: 45.9%, 20.3%, 18.4%, 13.2% and 6.6% respectively) (p<0.001).

**Figure 1.**
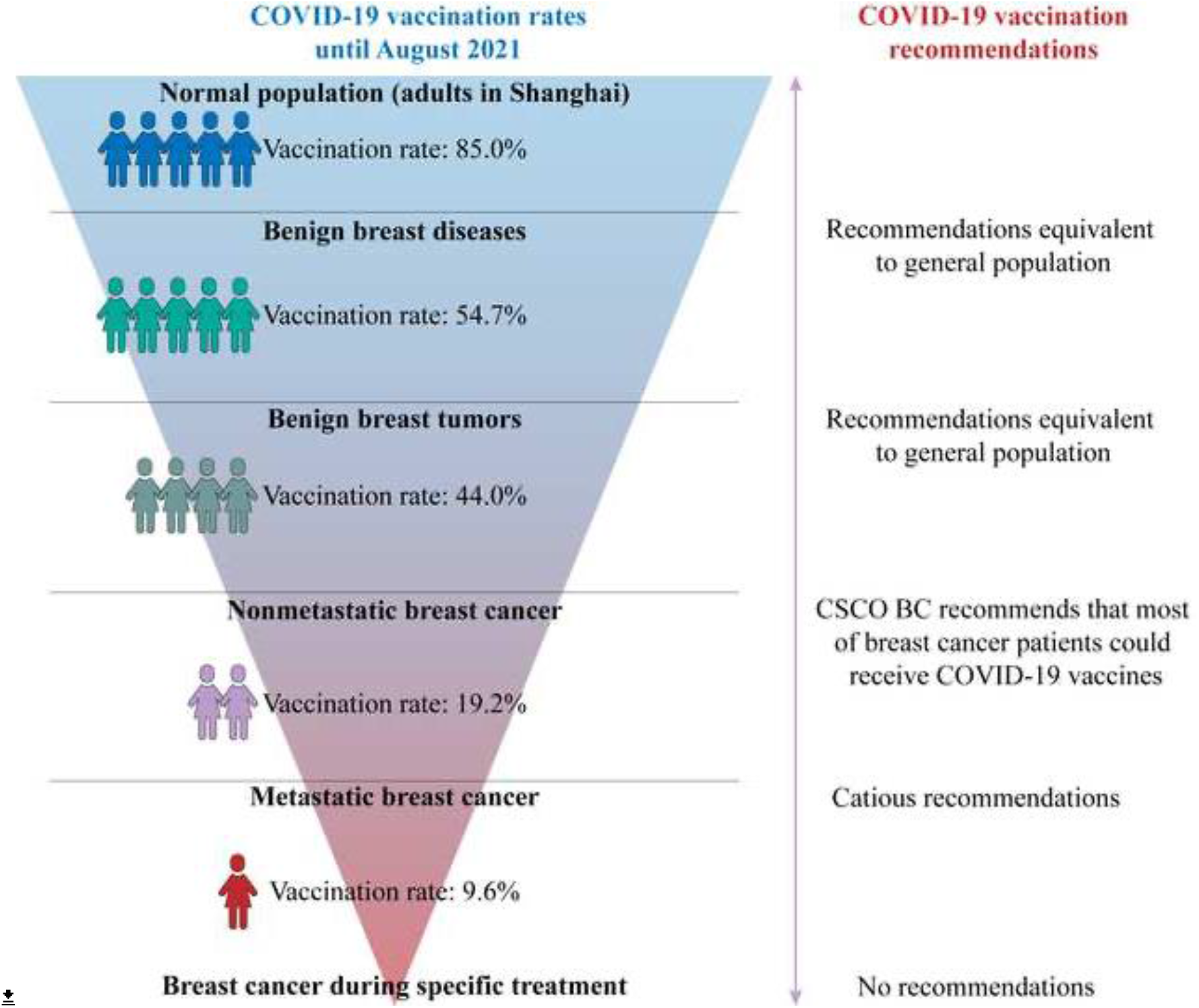
Vaccination rates and recommendations of vaccination in different groups of patients in August 2021. CSCO BC: The Chinese Society of Clinical Oncology-Breast Cancer.

**Table 1.** Clinical characteristics and vaccination rates of 11103 diagnosed breast cancer patients.

|  |  |  |  |  |  |  |  |
| --- | --- | --- | --- | --- | --- | --- | --- |
| cT stage | T0 (N=14) | Tis (N=283) | T1 (N=6418) | T2 (N=3876) | T3 (N=427) | T4 (N=85) | P value |
| Vaccinated | 1 (7.1%) | 130 (35.9%) | 1213 (18.9%) | 672 (17.3%) | 61 (14.3%) | 16 (18.8%) | < 0.001 |
| cN stage | cN0 (N=6886) |  | cN1 (N=2648) | cN2 (N=1089) | cN3 (N=475) |  |  |
| Vaccinated | 1449 (21.0%) |  | 454 (17.1%) | 134 (12.3%) | 55 (11.7%) |  | < 0.001 |
| cM stage | cM0 (N=10772) |  |  | cM1 (N=331) |  |  |  |
| Vaccinated | 2071 (19.2%) |  |  | 22 (6.6%) |  |  | < 0.001 |
| Clinical stage | Stage 0 (N=283) | Stage I (N=4759) | Stage II (N=4165) | Stage III (N=1565) | Stage IV (N=331) |  |  |
| Vaccinated | 130 (45.9%) | 968 (20.3%) | 767 (18.4%) | 206 (13.2%) | 331 (6.6%) |  | < 0.001 |

## Discussion

Our analysis observed the vaccination rates of patients with different breast diseases in one of the largest breast cancer centers in China. Our study found that breast cancer patients with a lower cTNM stage have a higher vaccination coverage rate, consistent with another research reported in June 2021 (12.63%).[6] In addition, the vaccination rate in patients with benign breast diseases was still lower than that of the general Chinese population until August 2021.

Notwithstanding that the proportion of the regional population vaccinated (viz. in Shanghai) has already surpassed the threshold for herd immunity (55.03% estimated in mainland China, and 85% estimated in adults in Shanghai in June 2021),[7, 8] the unvaccinated ones may still get infected via non-human sources, which may be a death blow for those with an immunocompromising state. Nevertheless, patients with benign breast diseases were granted with the equivalent recommendations of COVID-19 vaccines with general population, but the vaccination rate in these patients remained a low level of vaccination rate compared with that in general population. [9] In addition, on the one hand, the low vaccination rate in breast cancer patients could be explained by the impediment of specific adjuvant treatment, and misconception that COVID-19 vaccination is contraindicated in patients with breast cancer [6, 10]. Therefore, strict use of personal protective equipment is necessary in patients with advanced stage of breast cancers considering their intolerance for COVID-19 vaccines.

Moreover, the unexpected mutations of the virus leading to variant strains like Omicron and Delta have undermined the protection of herd immunity for virus spreading. Despite the lack of vaccine effectiveness data in cancer populations, the use of COVID-19 vaccines in immunocompromised populations shows low immunogenicity and reactogenicity, indicating that the vaccination itself won’t cause adverse outcomes to the patients. Therefore, on the other hand, breast cancer patients with lower clinical stage (viz., those in stage 0 and stage I) still hold partially competent immunity without receiving adjuvant chemotherapy or target therapy; hence vaccination can still induce potent active immunity and help reduce risks for adverse outcomes secondary to COVID-19 infection. In this sense, vaccination in BC patients is recommended, consistent with the Chinese expert consensus on COVID-19 vaccination for BC patients, unless patients were receiving several adjuvant treatments such as radiotherapy, chemotherapy, targeted therapy.[9]

This study has limitations, including its cross-sectional nature, reliance on self-reported data, and limited demographical data. Yet the study has enough data to shed light on the vaccination uptake in Chinese patients with benign breast diseases and breast cancer, suggesting a full-dose vaccination campaign for most of breast diseases patients (except for breast cancer during adjuvant treatment or in the advanced stage) tolerable for vaccination may protect them from unnecessary morbidity and mortality from SARS-CoV-2 infection. Since vaccination remains the leading strategy for preventing severe adverse outcomes. The active participation of oncologists is needed to further educate patients with breast diseases about the benefits and indications of COVID-19 vaccines, especially for those without vaccine-contraindication.

## Data Availability

All data produced in the present study are available upon reasonable request to the authors.

## Author Contributions

Dr Guang-yu Liu had full access to all data in the study and takes responsibility for the integrity of the data and the accuracy of the data analysis.

Concept and design: All authors.

Acquisition, analysis, or interpretation of data: Huai-liang Wu and Zong-lin He.

Drafting of the manuscript: Huai-liang Wu and Zong-lin He.

Critical revision of the manuscript for important intellectual content: All authors.

Statistical analysis: Huai-liang Wu, Zong-lin He, Yue Gong and Miao Mo.

Administrative, technical, or material support: Guang-yu Liu.

Supervision: Guang-yu Liu.

Huai-liang Wu and Zong-lin He contribute equally.

